# A population scale analysis of rare *SNCA* variation in the UK Biobank

**DOI:** 10.1101/2020.08.11.20172866

**Authors:** Cornelis Blauwendraat, Mary B. Makarious, Hampton L. Leonard, Sara Bandres-Ciga, Hirotaka Iwaki, Mike A. Nalls, Alastair J Noyce, Andrew B. Singleton

## Abstract

Parkinson’s disease (PD) is a complex neurodegenerative disease with a variety of genetic and environmental factors contributing to disease. The *SNCA gene* encodes for the alpha-synuclein protein which plays a central role in PD, where aggregates of this protein are one of pathological hallmarks of disease. Rare point mutations and copy number gains of the *SNCA* gene have been shown to cause autosomal dominant PD and common DNA variants identified using GWAS are a moderate risk factor for PD. The UK Biobank is a large prospective study including ~500,000 individuals and has revolutionized human genetics. Here we assessed the frequency of *SNCA* variation in this cohort and identified 26 subjects carrying variants of interest including duplications (n=6), deletions (n=6) and large complex likely mosaic events (n=14). No known pathogenic missense variants were identified. None of these subjects were reported to be a PD case, although it is possible that these individuals may develop PD at a later age, and whilst three had known prodromal features, these did not meet defined thresholds for being considered ‘prodromal’ cases. Four of the 14 large complex carriers showed a history of blood based cancer. Overall, we identified copy number variants in the *SNCA* region in a large population based cohort without reported PD phenotype and symptoms. Putative mosaicism of the *SNCA* gene was identified, however it is unclear whether it is associated with Parkinson's disease. These individuals are potential candidates for further investigation by performing *SNCA* RNA and protein expression studies, as well as promising clinical trial candidates to understand how duplication carriers potentially escape PD.

## Introduction

Parkinson’s disease (PD) often has a genetic component, either via rare damaging variants, more common risk variants, or a complex combination of these. Rare missense variants and copy number gains of the *SNCA* gene have been shown to cause autosomal dominant PD (Singleton et al. 2003; Polymeropoulos et al. 1997; Chartier-Harlin et al. 2004). Additionally, several non-coding common variants have been identified to moderately increase risk for PD (Nalls et al. 2019) and result in earlier onset (Blauwendraat et al. 2019) with a likely disease mechanism of increasing expression of SNCA (Pihlstrøm et al. 2018; Soldner et al. 2016). Importantly, *SNCA* encodes for the alpha-synuclein protein and aggregates of this protein have been identified in Lewy bodies, which is one of the pathological hallmarks of PD. Overall, this puts *SNCA* at a central position in PD pathogenesis and currently drugs are being developed targeting SNCA level lowering drugs.

Pathogenic missense *SNCA* variants are very rare in the general population with 5 disease-causing mutations (A30P, E46K, G51D, A53T and A53G). *SNCA* copy number variants appear to be more frequent compared to pathogenic missense variants, although still rare, with ~60 families reported. More rapid progression and atypical PD presentation is fairly common for all *SNCA* mutation carriers, with increased prevalence of symptoms like: dementia, rapid eye movement sleep behavior disorder, and autonomic dysfunction. Age of onset for *SNCA* missense variant and copy number variant carriers is earlier compared to idiopathic PD, with a broad range of age of onset in which late forties/early fifties tend to be the most frequent (Book et al. 2018; Konno et al. 2016).

Besides these autosomal dominant inheritance patterns, *SNCA* copy number gains also have been reported via mosaic patterns (Perandones et al. 2014; Perez-Rodriguez et al. 2019; Mokretar et al. 2018), although larger studies are needed to confirm whether this is a general PD pathogenesis mechanism. Here, we assess the presence and frequency of pathogenic *SNCA* variants (single nucleotide variants and copy number variants) in the population scale UK Biobank cohort (Bycroft et al. 2018).

## Methods

### UK Biobank genotype data

UK Biobank B allele frequency (BAF) and Log2 Ratio (L2R) were downloaded from the UK Biobank (v2) (June 2020) containing data of 488,377 individuals. BAF and L2R values were extracted for the larger *SNCA* gene region (chr4:84992449-96412248, hg19). BAF and L2R values were investigated in three *SNCA* regions 1) larger *SNCA* region gene +/-5 Mb (chr4:85645250-95759466, hg19), 2) *SNCA* gene +/-0.5 Mb (chr4:90145250-91259466, hg19), 3) *SNCA* gene body (chr4:90645250-90759466, hg19). For additional wider chromosome exploration the *SNCA* region gene, +/-20 Mb (chr4:70645250-110759466, hg19) and the full chromosome 4 region was used (chr4:1-180915260, hg19). For samples with large genomic events, all other autosomes were inspected as well. Typically BAF values of variants in the ~0.66 and ~0.33 range correspond to potential duplications. The number of BAF values of variants between <0.85 and >0.65 + <0.35 and >0.15 were counted for each of the three regions and if this variant count was higher than 6, the sample was plotted and visually inspected. L2R values were averaged for each of the three regions, where high average values would correspond to potential duplications and low average values would correspond to a potential deletion. After subject of interest selection, 363 were manually inspected of which 26 showed an event of interest including potential duplications, deletions and more complex events. BAF and L2R values were plotted in R (https://www.r-project.org/) using plot function and averages were calculated using the LOWESS (Locally Weighted Scatterplot Smoothing) function with variable the smoother span based on size of total graph (ranging from 0.01 to 0.0001). UK Biobank genotype data (v2) was used for calculating relatedness using PLINK (v1.9) (Chang et al. 2015).

### Exome Sequencing data

UK Biobank exome sequencing data (FE dataset, field codes: 23160 and 23161) were downloaded from the UK Biobank (June 2020) containing data of 49,960 individuals. Variants were annotated using ANNOVAR (Wang, Li, and Hakonarson 2010). Presence of known pathogenic variants were screened (A30P, E46K, G51D, A53E and A53T), reported pathogenic from ClinVar (https://www.ncbi.nlm.nih.gov/clinvar/) and HGMD variants (Professional 2020.2, Qiagen) and loss-of-function variants (stop, frameshift, splicing). Exome sequencing was used as replication of genomic events using the allelic depth (AD) field from the individual’s VCF files. AD ratio was calculated by dividing the highest depth value by the lowest depth value and then plotted using ggplot2 in R (Wickham 2016). Ten random samples were selected as negative controls. Additional population allele frequencies were obtained from the gnomAD (https://gnomad.broadinstitute.org/, v2.1.1, June 2020) (Karczewski et al. 2020).

### UK Biobank phenotype data

UK Biobank phenotype data was obtained from ICD10 codes (field code: 41270), PD (field code: 131023), illnesses of father and mother (field codes: 20107 and 20110), parkinsonism (field code: 42031) or dementia (field code: 42018), genetic ethnic grouping (field code: 22006), year of birth (field code: 34) and age of recruitment (field code: 21022). The MDS Prodromal Criteria were used to assess a potential PD prodromal phenotype (Heinzel et al. 2019).

### Data and code availability

All data used here is publically available upon application from the UK Biobank (https://www.ukbiobank.ac.uk/) and gnomAD (https://gnomad.broadinstitute.org/). Code used for preprocessing and analyses is available on GitHub repository https://github.com/neurogenetics/UKbiobank_SNCA.

## Results

Here we assessed the presence and frequency of potentially damaging *SNCA* mutations in the UK Biobank population level cohort. None of the five known *SNCA* pathogenic variants (A30P, E46K, G51D, A53E and A53T) were identified in the UK Biobank exome sequencing data. Two *SNCA* variants with conflicting or unconfirmed pathogenicity were detected (Sup Table 1) and all with relatively similar frequencies as in the gnomAD database. None of the variant carriers was a reported PD case, however three showed a positive family history for PD. Thirteen subjects carried *SNCA* P117T and one of these had a parent with PD. Additionally, 28 subjects carried the *SNCA* H50Q variant of which two had a parent with PD. Note that none of these observations showed statistically significant results (p>0.1, 2×2 Fisher exact test).

Next we explored the presence and frequency of *SNCA* copy number variants exploring BAF and L2R values in the UK Biobank genotyping data. 26 subjects of interest were identified and categorized into three groups, duplication carriers (n=6), deletion carriers (n=6) and large “complex” events carriers (n=14). Large complex events carriers were subjects where the BAF and L2R values did not meet the criteria for a “small” duplication or deletion but rather showed evidence of a large mosaicism event (Sup Table 2). Six potential *SNCA* duplications were identified (Figure 1), of which all showed clear BAF and L2R changes implying a *SNCA* duplication. The average estimated size was ~2Mb and ranging from ~0.8Mb to ~6Mb. Six potential *SNCA* deletions were identified of which five were complete gene deletions and one likely partial gene deletion (covering transcription start site and first exons) but sufficient to likely result in haploinsufficiency (Figure 2). The average estimated size was ~0.76Mb and ranging from ~0.2Mb to ~1Mb. Besides the identification of relatively straightforward duplications and deletions, 14 complex events were identified (Sup. Figure 1). Inspection of the wider SNCA (+/- 20Mb) and the full chromosome 4, and all autosomes suggested that most events are likely mosaicism events mostly spanning the majority of the long-arm of chromosome 4 (4q11-4q35). No additional large events on other chromosomes were identified in these subjects. After closer inspection eleven were classified as large complex events with likely varying levels of mosaicism, of which two were likely mosaic deletions likely due to a uniparental disomy event (Sup. Figure 1).

**Figure 1:**
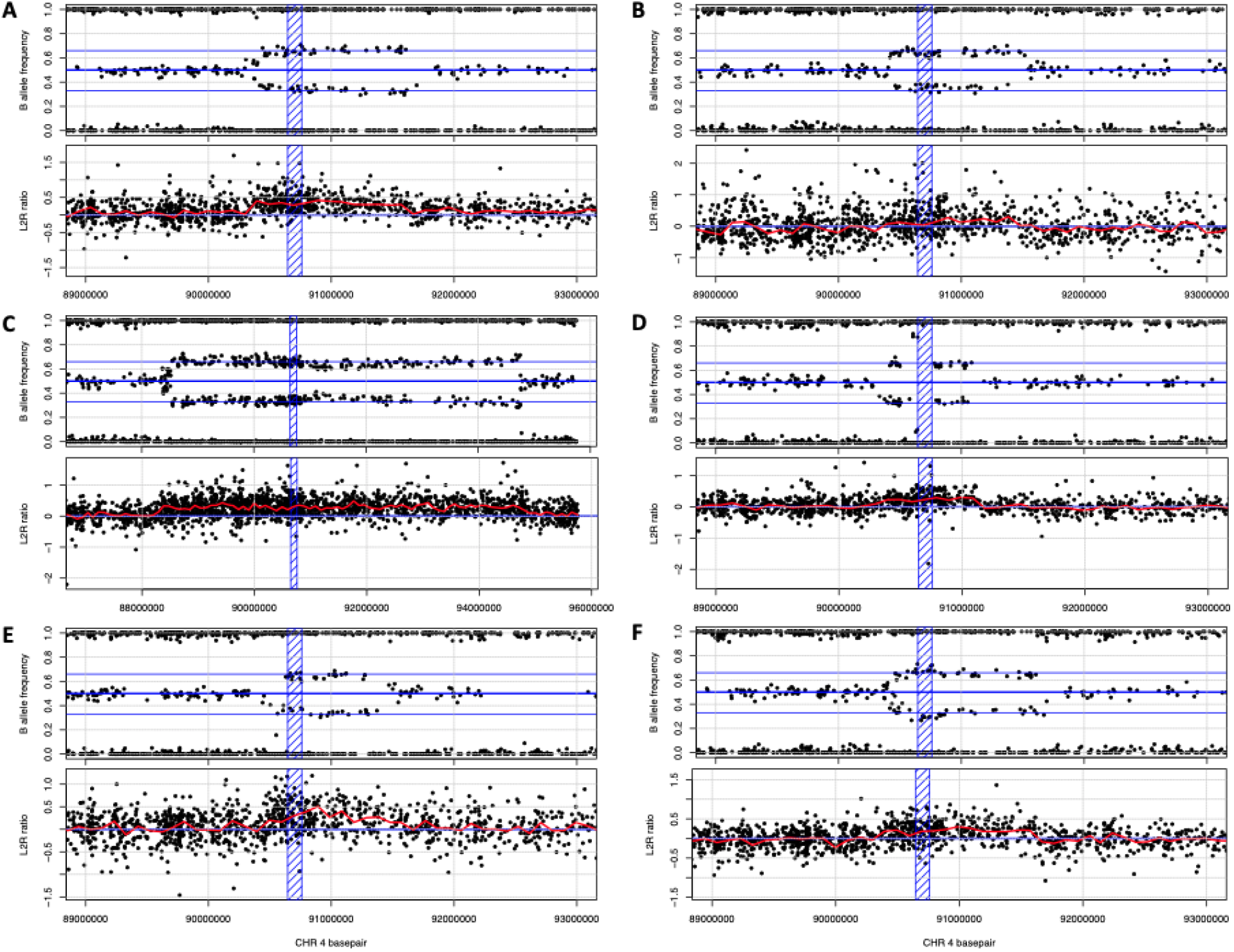
Six whole gene *SNCA* duplications were identified in the UK Biobank cohort.

**Figure 2:**
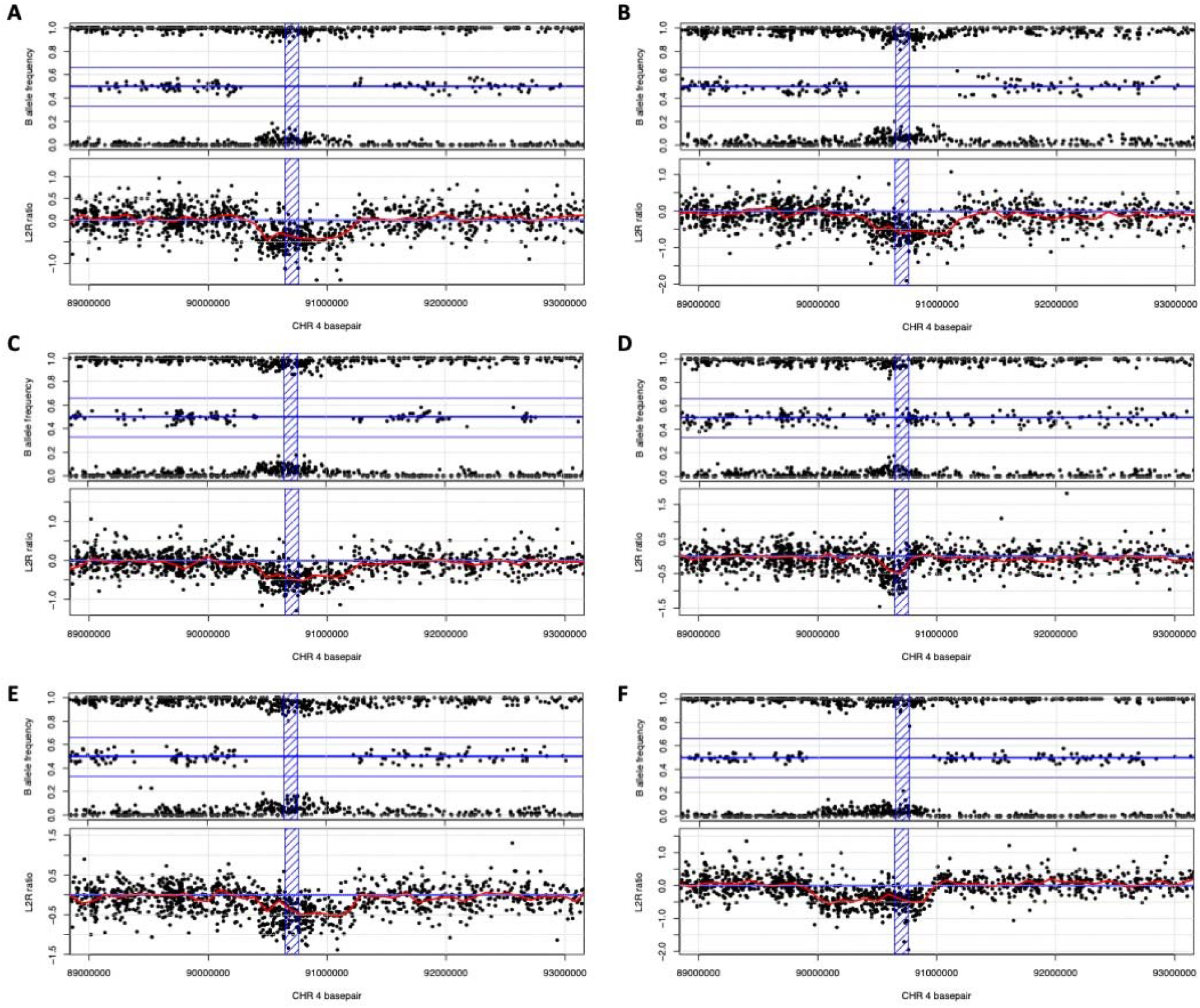
Five full *SNCA* deletions and one (likely) partial *SNCA* deletion (D) were identified in the UK Biobank cohort.

As partial validation of the results obtained from the genotyping array data, we explored exome sequencing allelic depth data (sequencing depth of each allele) of all heterozygous variants on chromosome 4. Exome sequencing was available for 2 of the 26 subjects with a potential *SNCA* alteration (subject #dup3 with *SNCA* duplication and subject #comp8 with *SNCA* complex). Subject #dup3 and subject #comp8 showed clear differences of allelic depth in the expected areas of genomic events (Figure 3). As negative controls we used 10 random subjects from the exome sequencing data of which none showed evidence of an allelic imbalance (Sup. Figure 2).

**Figure 3:**
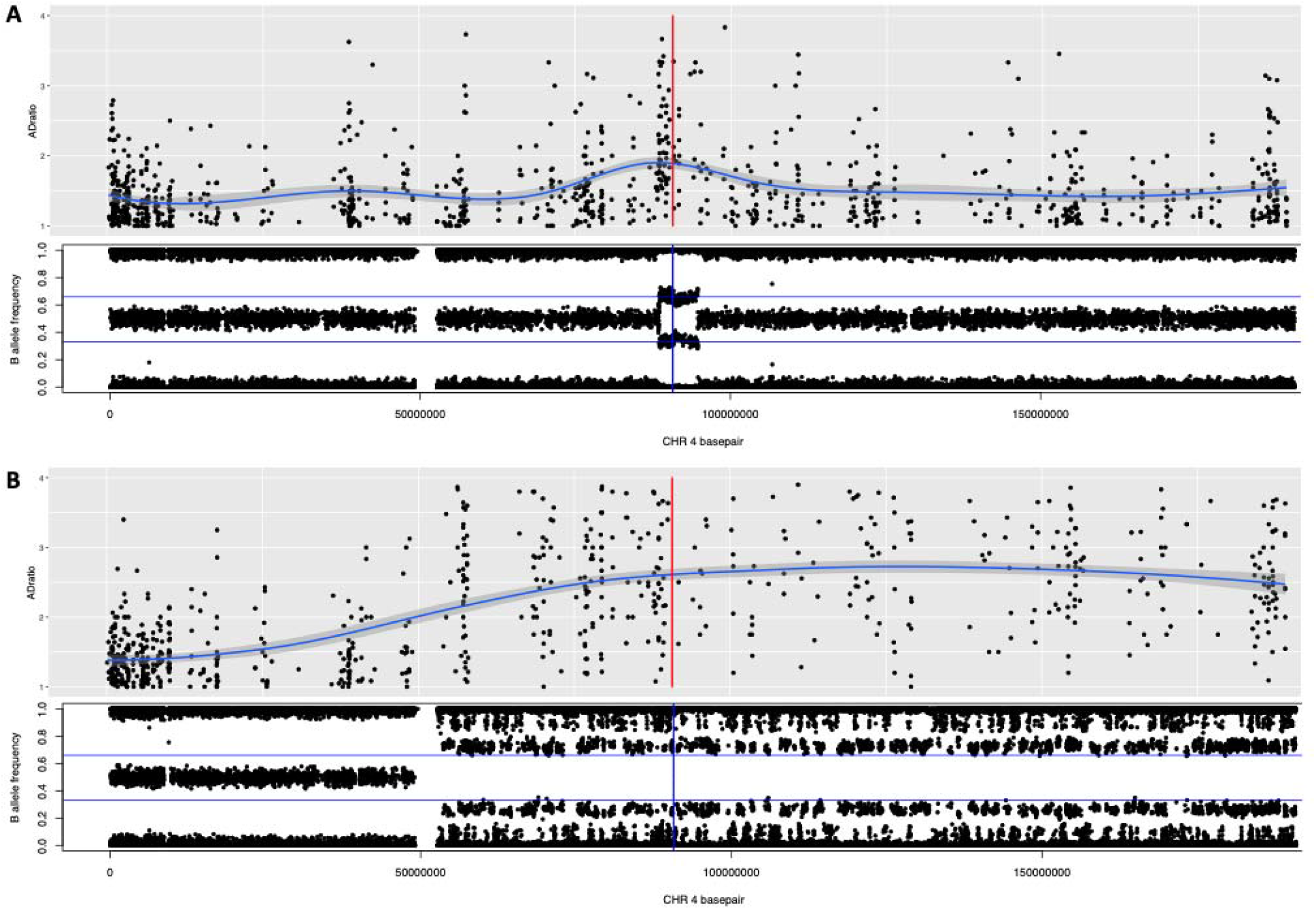
Partial validation of genomic events of the genotyping array data using exome sequencing data. A) Duplication in the *SNCA* gene region (subject #dup3), B) Partial duplication of the long arm of chromosome 4 (subject #comp8). Red and blue vertical line represents the SNCA gene body.

When assessing the UK Biobank phenotypic data of these subjects with a potential *SNCA* alteration, none of these subjects had been reported to be affected with PD, parkinsonism or dementia. Inspection of ICD10 codes also did not result in a potential PD diagnosis, although we cannot discard the possibility that some may be prodromal PD cases based on non-specific symptoms which are common in the older general population, including depression, anxiety, syncope, and head injury. Three participants were further evaluated to ascertain the probability of them being prodromal cases according to the recently revised MDS Prodromal Criteria. None of the participants met the threshold for probable prodromal PD (see Sup. Table 3). Of the 24 subject of interest, two had a parent with dementia (#comp3 and #comp6) and one had a parent with PD (#comp6). Note that none of these observations showed statistically significant results (p>0.1, Chi-square test). Four carriers (all large complex events) of the 14 (29%) potential mosaic subjects showed a history of blood-based cancer (Including: lymphoma, multiple myeloma, myelodysplastic syndrome, Sup. Table 2). Average age of recruitment of *SNCA* deletion carriers was 52 (range 41-56), for *SNCA* duplication carriers average age of recruitment was 61 (range 49-69) (all European) and for *SNCA* complex event carriers average age was 61 (range 42-69) of which two were not grouped as European. Interestingly, two pairs of individuals are siblings based on their relatedness value (PIHAT ~0.5) with reported birth years having ~2 years difference. Pair one (subject #dup1 and #dup6) carry both a 1.5 Mb *SNCA* duplication, and pair two (subject #del2 and #del5) carry both a 0.8 Mb *SNCA* deletion.

## Discussion

*SNCA* missense and copy number gains are a clear cause of autosomal dominant PD. Here we assessed the UK Biobank cohort for potential pathogenic variants in *SNCA*. None of the five known missense mutations were identified. Several other *SNCA* missense were identified, some with conflicting pathogenicity reports but none of these variant carriers was reported as a PD case. Three did report a positive family history for PD, however the allele frequency of these variants is too low to robustly test whether this frequency is more than expected by chance. Note that previously we suggested that *SNCA* H50Q is likely not pathogenic despite the presence of functional evidence (Blauwendraat et al. 2018).

We identified 6 copy number gains, 6 copy number loss and 14 complex *SNCA* alleles. Surprisingly none of these carriers have been reported as PD cases and although three had ICD codes that relate to recognised prodromal PD symptoms, none met published probability thresholds to be considered a prodromal case (Heinzel et al. 2019). Separately, one *SNCA* deletion carrier had an ICD code for schizophrenia which has been suggested to be a prodromal symptom in *SNCA* duplication carriers (Takamura et al. 2016). However, while *SNCA* copy number gains are a clear cause of autosomal dominant PD, *SNCA* copy number loss variants are to our knowledge not reported to cause PD. One possible explanation for the lack of PD status in these subjects is that they will develop PD at a later stage. The age of recruitment for the six duplication carriers is 61 (range 49-69). In a large recent meta-analysis of duplication carriers average age of onset was 46.9 with a range of 30-73 (Book et al. 2018) therefore it is possible that these subjects will develop PD at a later age.

The identification of 14 potential mosaic events is of interest, especially given the prior evidence of *SNCA* mosaicism in PD and synucleinopathies (Perez-Rodriguez et al. 2019; Mokretar et al. 2018; Perandones et al. 2014). Eleven were classified as large complex events with likely varying levels of mosaicism (Conlin et al. 2010). Two were likely mosaic deletions of the larger *SNCA* region with 20-25Mb size, while one was likely a uniparental disomy event spanning the majority of the long arm of chromosome 4. In a subset of the UK Biobank data, mosaic effects have already been shown to be present on a larger scale than previously assumed (Loh et al. 2018). Additionally, as expected, four (all large complex events) of the 14 (29%) potential mosaic subjects showed a history of blood-based cancer. It is important to note that the current data is based on blood derived DNA, and it is unclear whether these effects are also present in more PD related tissues and cells like the brain and neurons. Besides it is unclear what the effect of these events is on *SNCA* gene expression and a-synuclein protein expression. A deeper investigation of these genomic events is needed before robust disease associations can be made.

Although our findings are based on a population level cohort there are several clear limitations. First, the gold standard of copy number variant detection is multiplex ligation-dependent probe amplification (MLPA). However, DNA of these subjects is not readily available making validation of these genomic events not possible. In an attempt to overcome this limitation, we partially validated two genomic events using the available exome sequencing data in which clear events were detected, although gene and protein expression investigations would be necessary to verify the functional consequences. More future extensive validation is needed to determine the exact size and downstream effects of these variants especially of the complex mosaic events. Second, although the UK Biobank is the largest and most comprehensive genotype/phenotype cohort to date there is the potential of missing phenotype data when using electronic health record (EHR) ICD10 codes for various reasons. Therefore PD or a prodromal PD phenotype could be missed in some instances however being unlikely that this information is missing in all carriers of potential duplication events. Third, our approach is partly based on manual interpretation of prioritized subjects and is biased due to the genotyping array design (spacing and number of variants in the *SNCA* gene region) toward larger events and smaller deletions (eg. single exon) or complex rearrangements are easily missed. Although manual interpretation is not ideal it is more likely to be conservative than automatically generated algorithms since these often also require a manual interpretation step. Of note, multiple copy number variant events were detected outside of the *SNCA* gene body region which can have a significant impact on expression. To confirm this, RNA sequencing data would be needed.

Overall, here we identified that copy number variants and mosaicism in the *SNCA* region are present in the general population without reported PD symptoms. These subjects are outstanding candidates for more thorough investigation for *SNCA* levels, general *SNCA* biology, potential clinical trial candidates and how these duplication carriers potentially escape PD. Mosaicism of the *SNCA* gene region in blood derived genotype data is of interest but this mechanism needs to be explored on a larger case-control scale to assess whether this is associated with disease or not.

## Data Availability

All data used here is publically available upon application from the UK Biobank (https://www.ukbiobank.ac.uk/)

## Acknowledgements and Funding

We would like to thank all of the subjects who donated their time and biological samples to be part of this study. This research was supported in part by the Intramural Research Program of the NIH, National institute on Aging. This research has been conducted using the UK Biobank Resource under Application Number 33601. This study used the high-performance computational capabilities of the Biowulf Linux cluster at the National Institutes of Health (http://hpc.nih.gov).

